# Development of the Sm14/GLA-SE - schistosomiasis vaccine candidate: An open, not placebo-controlled, standardized dose immunization phase Ib clinical trial targeting healthy young women

**DOI:** 10.1101/2022.08.17.22278904

**Authors:** Marilia Santini-Oliveira, Patrícia Machado Pinto, Tatiane dos Santos, Monica Magno Vilar, Beatriz Grinsztejn, Valdilea Veloso, Elan C. Paes-de-Almeida, Maria A. Z. Amaral, Celso R. Ramos, Miryam Marroquin-Quelopana, Rhea Coler, Steven Reed, Marcia Ciol, Wilson Savino, Juçara de Carvalho Parra, Marília Sirianni dos Santos Almeida, Miriam Tendler

**Affiliations:** Evando Chagas National Institute of Infectology, Fiocruz, Rio de Janeiro, 21045-900, Brazil; Laboratory of Experimental Schistosomiasis, Oswaldo Cruz Institute, Oswaldo Cruz Foundation (Fiocruz), Av. Brasil, n°4365, Manguinhos, 21045-900, Rio de Janeiro, Brazil; Department of Basic Sciences, Fluminense Federal University, Nova Friburgo, RJ, Brazil; Program for Development of the Mata Atlântica Campus, Fiocruz, Rio de Janeiro, Brazil; Fenix Biotec Treinamento SS LTDA, São Paulo, Brazil; Department of Global Health, University of Washington, Seattle, USA; HDT Bio, Seattle, USA; Department of Rehabilitation Medicine, School of Medicine, University of Washington, Seattle, USA; Laboratory on Thymus Research, Oswaldo Cruz Institute, Oswaldo Cruz Foundation, Rio de Janeiro, Brazil; Brazilian National Institute of Science and Technology on Neuroimmunomodulation, Oswaldo Cruz Institute, Oswaldo Cruz Foundation, Rio de Janeiro, Brazil; René Rachou Institute — Fiocruz Minas, Oswaldo Cruz Foundation, Belo Horizonte, Brazil

**Keywords:** Vaccine, Schistosomiasis, Phase Ib clinical trial, toxicology, pregnant rabbits, Sm14 protein

## Abstract

We report the successful closure of Phase I clinical trials of the *Schistosoma mansoni* 14 kDa fatty acid-binding protein (Sm14) + glucopyranosyl lipid A in squalene emulsion (GLA-SE) vaccine candidate against human Schistosomiasis, comprising Phases Ia and Ib. Shown here are the results of Phase Ib, an open, not placebo-controlled, standardized-dose immunization trial, involving 10 healthy 18-49 years old women submitted to the same clinical protocol and the same batch of cGMP Sm14+GLA-SE used in Phase Ia, which was one on men. Fifty µg Sm14 protein plus 10 µg GLA-SE per dose were given intramuscularly thrice at 30-day intervals. Participants were assessed clinically, biochemically, and immunologically for up to 120 days. In preambular experiments involving vaccinated pregnant female rabbits, we did not find any toxicological feature either in offspring or mothers, as ascertained by histopathology and biochemical parameters. The vaccine induced adaptive immunity in the animals, as defined by the detection of anti-Sm14 antibodies in the sera. In women, neither serious nor light adverse events were observed. Sm14+GLA-SE vaccination induced high titers of anti-Sm14 serum IgG antibody production. Total anti-IgG serum levels remained high 120 days after the first vaccination dose. Significant increases in Sm14-specific total IgG, IgG1, and IgG3 were observed 30 days after the first vaccination, with specific IgG2 and IgG4 after 60 days. Sm14+GLA-SE vaccination also elicited robust cytokine responses with increased TNFα, IFNγ, and IL-2 profiles in all female vaccinees on days 90 and 120.

As in Phase Ia, the Sm14+GLA-SE vaccine was shown to be strongly immunogenic and well tolerated. The completion of Phase I clinical trials performed to the highest standards set by the Good Clinical Research Practice (GCP) standards and pre-clinical data in pregnant rabbits enabled the vaccine candidate to proceed to Phase II clinical trials in endemic areas.

**Study registration ID:** NCT01154049 at http://www.clinicaltrials.gov. Brazilian Registry of Clinical Trials UTN: U1111-1135-6815

## Introduction

Schistosomiasis is a parasitic disease that affects populations living in areas with very poor sanitary conditions where they get infected through the contact with contaminated water during their daily working, domestic or leisure activities. Children are the main target of the infection that strongly compromises their physical and cognitive development [1,2]. Such a feature turns schistosomiasis, not only into a disease resulting from poverty, but also into a backwardness generator in endemic countries. Schistosomiasis is present in tropical areas of America, Medium East, Asia and Africa, where the impact over public health is catastrophic as a consequence of the size of the territory affected as well as the number of people getting infected. The World Health Organization (WHO) estimates that over 240 million people living in 78 countries are infected and around 800 million live in areas with risk of transmission and infection and are part of global programs directed to the effective control of transmission of schistosomiasis [3]. According to the Ministry of Health, in Brazil, around 1.5 million people live in areas of risk of acquiring the disease [4]. Yet, we still lack a vaccine for protecting the populations from the disease.

Presently, the so-called Mass Drug Administration (MDA) program against schistosomiasis is the strategy adopted by WHO in which populations in endemic areas are treated annually with Praziquantel without previous diagnosis. Such strategy actually led to an improvement with regards to the pathology associated to schistosomiasis [5]. However, the world prevalence is still as high as it has always been and the Disability-Adjusted Life Years (DALYs) value, which is an important tool to assess the impact of diseases, has increased [6].

The Sm14 vaccine candidate has been developed at the Oswaldo Cruz Foundation (Fundação Oswaldo Cruz, Fiocruz, Brazil) as a humanitarian vaccine to be used on the population living in areas with risk of infection [7–9].

The Sm14 recombinant protein is currently the basis for a molecular antiparasitic vaccine associated to GLA-SE adjuvant has been developed as an anthelmintic bivalent vaccine, directed against fasciolosis and schistosomiasis. Sm14 is a member of the Fatty Acid Binding Proteins family, originally isolated from *S. mansoni* [10], already identified in most parasite helminths of human and veterinary importance [5–15].

During the experimental phase of Sm14 vaccine development, concentrated in vaccination of outbred animals, it was possible to demonstrate consistent protection against challenge infection with cercariae of *S. mansoni* and significant reduction of adult worm burdens of vaccinated/infected animals evidenced by mean values of adult worm loads and additional measures of the distribution of frequencies of Swiss-Webster mice in sequential worm burdens rates [16–19]. Therefore, one can expect that vaccination with Sm14+GLA-SE in human may lead to the reduction of worm burdens, reducing reinfection rates and diminishing the aggressive inflammatory response that has been observed following interrupted chemotherapy in children living in high-transmission areas [20,21].

The major objective of the Sm14 vaccine project is to promote health access through a safe and immunogenic vaccine capable of blocking the transmission of the disease. This will avoid that populations continue to receive repeated doses of chemotherapeutic drugs during childhood and youth. The design of the vaccine foresees its large-scale use associated to the other control measures already known such as chemotherapy with Praziquantel, sanitation, health education, etc., capable of promoting the control of the disease and mitigating the suffering historically produced by this disease in tropical countries.

The Brazilian National Sanitary Surveillance Agency (Anvisa) approved the start of the Clinical Protocol for Phase Ia Study (CE# 990768/10-8), nonendemic area of Rio de Janeiro, Brazil, with 20 healthy male volunteers, which was completed with success and validated. The validation was done with the use of a Data Bank fed with all records of the study for calculations and detailed and descriptive statistical analyses of results [22]. In order to comply with the requirements for a similar study in women, Anvisa requested experiments done in pregnant experimental animals. On a second round, and after completion of a requirement for testing the toxicity of the product under investigation, Anvisa approved the Phase Ib Clinical Trial, same Brazilian region as the phase 1a clinical trial, this time with female volunteers. As required by the regulatory agency, the Sm14+GLA-SE formulation was preambularly tested and successfully approved in pregnant rabbits.

The step related to process of vaccination on Phase Ib (with young adult female volunteers) was conducted by the Evandro Chagas National Institute of Infectology (INI/Fiocruz) using the same batches of the Sm14+GLA-SE used on Phase Ia.

The immunogenicity induced by the vaccine Sm14+GLA-SE in women was also approached by means of a platform made available by the Access to Advanced Health Institute (AAHI) (previously Infectious Disease Research Institute IDRI) Seattle, USA to where sera and cells collected in different points of the study from volunteers were shipped. The steps needed to attest safety and immunogenicity of the vaccine and validation of the study were completed and are described in the present work.

## Materials and Methods

### Vaccine Antigen and Adjuvant

For the studies carried out in rabbits as well as the clinical trial, the Sm14 vaccine used in the study was produced at the Ludwig Institute for Cancer Research (LICR) and Cornell University, NY, USA under Good Manufacturing Practice conditions (cGMP). Hermetically closed vials, with individual doses (0.55 mL), labelled with product name, Sm 14, lot number (PBR-0057-002), date of manufacture and “*only for investigational use*”, were shipped to Brazil under the import authorization from the National Council for Scientific and Technological Development (CNPq) and the Anvisa. Vials were kept at 2-8°C under continuous temperature control from the origin at Fiocruz (Rio de Janeiro) until use, with regular inspections assuring contents remaining liquid and transparent without suspended particles.

The Infectious Diseases Research Institute/ Access to Advanced Health Institute (IDRI/AAHI) provided the Glucopyranosyl Lipid-A adjuvant in Squalene Emulsion (GLA-SE) vials containing 0.4 mL, at a concentration of 40 µg/mL, also produced under GMP conditions, labelled with lot number (Part # 0037-09M001) and manufacturing date. Shipping and storage followed the same procedures as described above.

The batches of Sm14 and adjuvant were filled finished as single doses and mixed at the moment of immunizations.

To formulate the vaccine antigen dose at the time of administration, 0.4 mL of a 200µg/mL Sm14 suspension were aspirated using a 1 mL sterile syringe. Subsequently, the volume of the syringe was injected into the vial containing the GLA-SE adjuvant, obtaining a total volume of 0.8 ml mixture. After mixing by inverting the vial, 0.5 mL containing 50µg of Sm14 protein and 10µg adjuvant was administered by intra-muscular route to both animals and women volunteers. The dose used in these studies were based on a clinical protocol approved by Anvisa for the Phase Ia study in male volunteers [22]. The two components of the vaccine candidate, kept in separate vials, were monitored by Pharmaceutical Product Development (PPD) [23]. Samples were sent to PPD for control every three months in accordance with applicable standard operating procedures and the US Food and Drug Administration (FDA) regulations. Stability of the Sm14 GMP lot was followed by PPD and updated “Certificate of Analysis” issued.

### Pre-clinical experiments on safety and immunogenicity of pregnant rabbits and respective off springs, following Sm14 vaccination in the mothers

The study in pregnant rabbits was conducted in accordance with the protocol provided by Anvisa. It was reviewed and approved by the Ethics Committee on Animal Research of Fiocruz (CEUA/Fiocruz) n° L0063/08 – LW 19/2013.

The reproductive toxicity tests were performed with the same vaccine lot further used in Phase I Clinical Trials, which was produced under current good manufacturing practice (cGMP) conditions by LICR/Cornell University, NY, USA. Vials containing individual doses (0.55 mL, 200 µg/mL) were labelled with the product name (Sm14) and lot number (PBR-0057-002). Vials were kept at +2-8°C under continuous temperature control until use, with regular inspections to assure that the contents remained liquid and transparent without suspended particles.

Twenty-four NZW pregnant rabbits, with the same age and weight at the beginning of the experiment, were divided into two groups just after mating: one group comprising 12 rabbits immunized with three equal doses of 50 µg Sm14+10 µg GLA-SE/dose, with a 7-day interval between doses; and the control group, comprising 12 rabbits injected with PBS, same days intramuscularly. Injections of 0.5 mL/dose for each dose schedule were at the external middle third of the right and left thigh alternately, and administered on days 0 (day 9 of pregnancy), 7 (day 16 of pregnancy), and 14 (day 23 of pregnancy). The pregnancy period of these animals is approximately 30 days. Necropsy and anatomopathological evaluations were performed on day 29 of pregnancy.

All animals had the inoculation area shaved and marked before each immunizing dose, to assess further possible local reactions that might include erythema and edema; both being scored according to standard protocols [24]. Daily clinical evaluation of each animal was performed and duly recorded in the individual data chart by a veterinarian. Parameters such as occurrence of abortion, stress level, appetite, manure, and skin/local reactions at inoculations sites, were evaluated and measured. Body weight was determined at the beginning and at the end of the study. Food and water consumption was recorded every day individually.

Biochemical and blood cell evaluation was done following current laboratory practices and included liver, renal and pancreatic functions, as well as red and white blood cell countings and coagulation parameters.

All animals were anesthetized with thiopental sodium and euthanized by exsanguination. Detailed anatomopathological analyses of all organs were carried out following euthanasia. All animals were subjected to a macroscopic inspection of the organs and cavities. Samples of organs were collected in buffered 10% formalin for later microscopic analysis. The ovaries, uteri, and fetuses were collected and fixed for further analysis, such as number, size, weight, *corpora lutea*, placenta and living fetuses. Paraffin-embedded tissue sections were stained with hematoxylin and eosin (H&E) and examined under a light microscope [25].

Serum samples were collected from all rabbits before the start of pregnancy and on the date of euthanasia. The total anti-Sm14 IgG antibody titers in rabbit sera samples were determined by ELISA [26] in accordance with previously described procedures [27]. Briefly, the wells were coated overnight at 4°C with 100 µL/well of Sm14 protein solution at 4 µg/mL in carbonate buffer 0.1 M pH 9.6, blocked for 1h at 37°C, and then serum samples were added in two-fold serial dilution from 1/100 up to 1/1600. After washing, peroxidase conjugated goat anti-rabbit total IgG specific to the gamma chain (Santa Cruz Biotechnology, USA) was added to each well, and the plates were incubated for 1h at 37°C. Wells were washed and the reaction was developed by the addition of TMB substrate (Sigma-Aldrich, T0440) for 15 min at room temperature in the dark followed by the addition of Stop reagent (Sigma-Aldrich S5814); subsequently, the OD at 450 nm was measured using a BIORAD device, Model 680.

All statistical analyses were carried out by using GraphPad Prism version 4.0 software. All analyses were conducted by applying the two-tailed test, with *p*-values below 0.05 considered to be significant.

### Phase Ib clinical trial in healthy women

The methodology used to conduct the Sm14+GLA-SE candidate vaccine Phase 1b clinical trial in women was essentially the same described by Santini-Oliveira et al. [22] for Phase 1a with healthy male adults (CE # 990768/10-8 - CAAE 0018.0.009.000-10). The protocol for the Phase Ib trial was approved by Anvisa (CE# 0577594139) and by the Evandro Chagas Review Council of the National Institute of Infectious Diseases (INI), Fiocruz (CAAE 09627212.4.0000-5262). In addition, tests were monitored by TECHTRIALS, an independent research company, and data was deposited into a Data Bank [28] under the code NCT01154049.

Twenty healthy female volunteers from a non-endemic area of Brazil (City of Rio de Janeiro) were enrolled and registered under numbers from #1 to #20. Among them, 10 were selected and went up to the end of the study. Inclusion criteria comprised negative pregnancy test, availability during the whole duration of the trial, no evidence of any acute infection, no previous or ongoing serious chronic disease, normal findings for organ functions as demonstrated by laboratory tests and physical examination of the cardio-respiratory and brain/neurological, abdominal organs, skin, and lymphatic system. Exclusion criteria included pregnancy/breast feeding, abnormal hematology or blood chemistry counting or data, any kind of ongoing infection, including infection by the human immunodeficiency virus (HIV), any chronic disease, alcohol/drug abuse or history of medication or vaccination in the preceding month. After clinical screening, the 10 participants aging 26-48 years (mean=36.3, sd=6.8) were selected for the trial, and their demographic characteristics are depicted in S1 Table.

Full physical examens and laboratory tests were done on baseline and days 0, 7, 30, 37, 60, 67, 90 and 120 (Supplemental figure 1). Participants were monitored at the site up to one hour after vaccination and followed-up by phone 20-28 hours afterwards. According to Santini-Oliveira [22], all adverse episodes were evaluated in compliance with the International Centers for Tropical Disease Research network (ICTDR), and according to the degree of severity as follows: Level 1 (tolerable with mild discomfort and not interfering with regular activities); Level 2 (discomfort with the impairment of regular activities but not requiring any specific medical assistance, or treatment); Level 3 (requiring discharge of regular activities and medical intervention) and Level 4 (life threatening). The laboratory at the Evandro Chagas National Institute of Infectology (INI) is certified by the College of American Pathologists (CAP, EUA) and laboratory tests were evaluated based on the INI Standard Values Chart. The study followed FDA regulations [29].

Samples of peripheral blood mononuclear cells (PBMCs) and of sera were collected from all subjects on baseline and days 0, 30, 60, 90 and 120. Immediately after collection, PBMCs were separated by centrifugation at room temperature in Ficoll-Paque Plus (GE Healthcare, USA) and mixed with Fetal Bovine Serum (FBS) (Gibco Life Technologies, USA) and 10% DMSO. Cells were subsequently transferred to 2 mL cryovials and frozen down to −80°C in a NALGENE ™ Cryo 1°C freezing container (Nalgene, USA) overnight. On the following day vials were transferred to liquid nitrogen and later dry shipped to the Infectious Disease Research Institute (IDRI, Seattle, USA) where they were stored in liquid nitrogen until use.

Sera were also stored in Brazil at −80°C and transferred in dry ice to IDRI for analysis. At IDRI, sera were stored at −80°C, till experiments were performed. The immunogenicity analyses included isotype check of anti-Sm14 antibodies and cell-mediated responses specifically induced after vaccination with Sm14+GLA-SE. It also included detection and quantification of intracellular cytokines after stimulation of PBMCs with purified rSm14. Immune cell responses were measured using the PBMC Luminex Kit for T cell cytokines (Milliplex ®MAP) [22]. Isotyping and the correlation between the development and the size of humoral response measured in terms of IgG against the Sm14 protein were done by enzyme-linked immunosorbent assay ELISA [22,26].

Descriptive analysis using IBM SPSS v20 statistical package was performed for all variables. Boxplots for all follow-ups assessed whether observations were outside the limits specified for specific laboratory tests. Proportions of adverse events and 95% confidence intervals were calculated using Clopper-Pearson method [30]. For toxicity grading scales, we used limit values based on published information when available, which included clinical experience and reviewed vaccine clinical trials on healthy subjects. Toxicity grading scales were based on the local laboratory reference values when those were defined. Statistical analyses of immunogenicity data were performed using analysis of variance, followed by Tukey’s test for multiple comparisons.

The statistical analysis was done based on the evaluation of local and systemic reactogenicity observed in the first 36 hours after each dose of the vaccine. Results of chemical and hematological exams were compared to reference values in accordance with the Richet Laboratory (LR) criteria and ICTDR, including levels of toxicity as defined by FDA [29].

## Results

### Sm14 vaccine candidate does not induce any toxicological effect in pregnant rabbits

The body weights and structures of the analyzed organs in all pregnant rabbits, irrespective of being or not vaccinated were within normal limits, according to established criteria [31]. Macroscopic analysis of organs and cavities revealed results consistent with the usual aspects for normal pregnant rabbits [32]. The ovaries of all 24 animals were consistent with normal appearance. All analyzed uteri were comparable with a normal pregnant uterus for outbred rabbits according to established criteria [33]. The microscopic results showed that the ovaries of all 24 pregnant rabbits had follicles at different stages of development in the cortical region, usual interstitial stromal glands and containing varying numbers of *corpora lutea* (Figure 1). There was no statistically significant difference for any of the parameters assessed in this study between vaccinated and control animals. Table 1 shows that the duration of pregnancy, the number of implantation sites in the uterus, and the number and viability of pups guarantee the absence of maternal and embryo-fetal toxicity in pregnant rabbits vaccinated with Sm14+GLA-SE. The results were also consistent with the morphological and physiological aspects reported for farmed NZW rabbits [31–33].

**Table 1.** Sm14 vaccination does not induce changes in pregnancy parameters.

| Experimental Groups | Number of fetuses | Size of fetuses (cm) | Weight of Fetuses (g) | Number of corpora lutea | Number of Placentae (deployments) | Number of Live Fetuses | Percentage of Live Male Fetuses/Litter |
| --- | --- | --- | --- | --- | --- | --- | --- |
| Sm14+GLA-SE | 6.60 ± 2.78 | 6.85 ± 2.40 | 23.43 ± 11.32 | 6.33 ± 3.14 | 7.58 ± 3.02 | 83 | 56.25 ± 30.24 |
| PBS Control | 7.00 ± 3.24 | 7.10 ± 0.83 | 21.58 ± 7.32 | 7.25 ± 2.89 | 8.42 ± 2.99 | 84 | 55.60 ± 34.91 |

Data are presented as the mean ± standard deviation. The parameters were evaluated by Student's *t*-test to compare the mean values of groups. In all cases, differences were considered statistically significant for p values of < 0.05. No differences were found.

**Fig 1.**
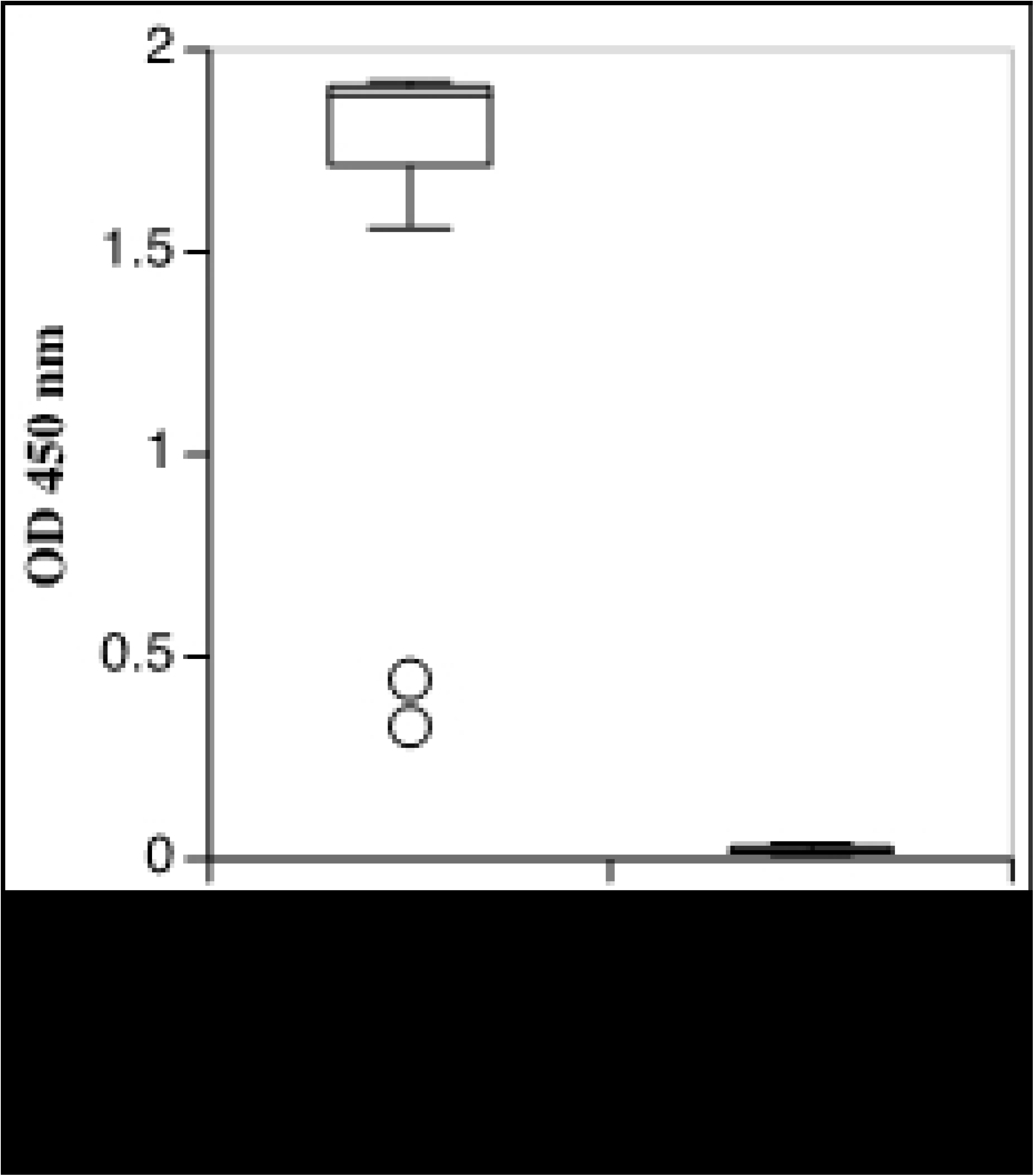
Normal macroscopic and microscopic views of an ovary from a Sm14/GLA-SE vaccinated pregnant rabbit. Panel **A** depicts the normal macroscopic pattern of the ovary, showing multinodular outer surface and brownish with marked vessels. Panel **B** shows microscopic view with follicles in various stages of development in the cortical region (within the black circles). Corpora lutea (CL) can be seen in panel **C** whereas a higher magnification of a follicle can be seen in Panel **D**. Sections were stained with Hematoxylin-eosin. Scale magnification with the objectives for 40x (in panel B), 100x (in panel C) and 200x in panel D).

The skin and subcutaneous tissue were removed from the injection sites of the immunized animals. The results of the individual tests showed no significant histopathological alterations in the skin at the immunization site. In the anatomopathological evaluation of the organs, it was observed that all groups, including the control group, had small and scarce foci of calcification in 5% of the cortical and medullary tubules of the kidneys.

Lastly, ELISA was used to analyze sera collected from the animals after the final injection and showed considerably high production of the anti-Sm14 antibody in the sera of vaccinated versus non-vaccinated PBS-treated pregnant rabbits. Anti-Sm14 antibody levels (evaluated by optical density) were clearly high in the vaccinated group as compared to controls (Figure 2).

**Figure 2.**
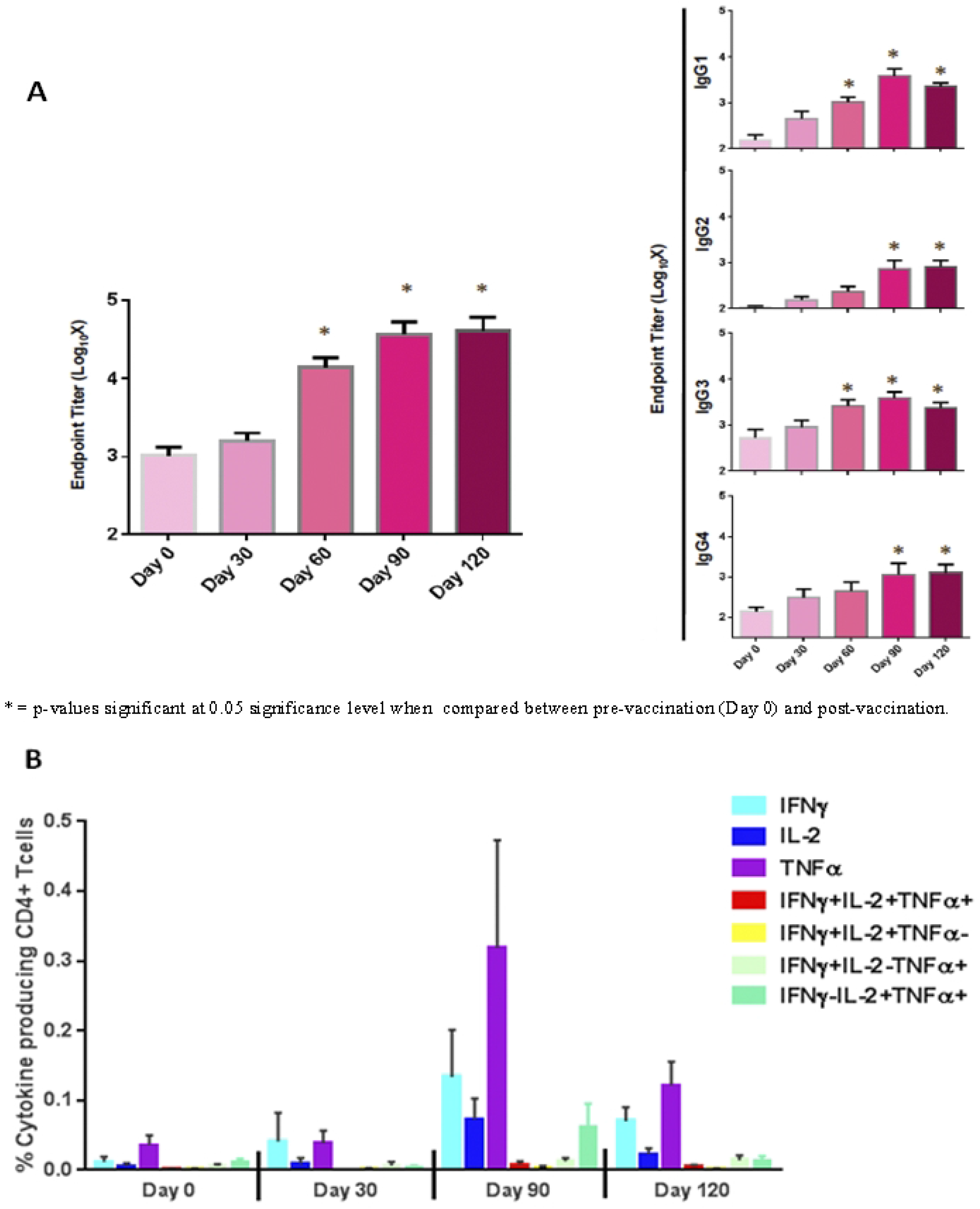
Total anti-Sm14 IgG antibody titers, determined by ELISA, of all rabbit serum samples from the two groups in the reproductive toxicity study. OD: optical density.

### Sm14 vaccine candidate does not induce any major adverse effects (AEs) in healthy women

The certification of Sm14+GLA-SE vaccine safety in this preclinical study paved the way for the phase Ib clinical trial of the vaccine in young adult non-pregnant women. The candidate vaccine Sm14+GLA-SE was shown to be safe with no serious adverse events as observed in the participants after any of three doses. No local reactions at the site of the injection occurred, except for local pain which affected 9 out of 10 women after the first dose, 5 out of 9 after the second, and 4 out of 10 after the third (all lasting less than 24 hours after injection). Only one subject reported involuntary muscle contractions lasting 5 minutes near the site of infection after the first dose. Table 2 shows the adverse events associated with vaccination of the healthy female volunteers in the Phase Ib.

**Table 2.** **Adverse events related to the injection of vaccine: reactions at the site of the injection and systemic reactions during the first 36 hours after the administration of the vaccine**

| Type of Adverse Effect | First Dose<br>(Day 1) | Proportion<br>(95% Confidence Intervall) |  |
| --- | --- | --- | --- |
|  |  | Second Dose<br>(Day 31) | Third Dose<br>(Day 61) |
| **Number of participants** | 10 | 10* | 10 |
| **Serious adverse effects** | 0.000<br>(0.000, 0.308) | 0.000<br>(0.000, 0.308) | 0.000<br>(0.000, 0.308) |
| **Reactions in the of the injection** |  |  |  |
| Reaction to touch | 0.000<br>(0.000, 0.308) | 0.000<br>(0.000, 0.308) | 0.000<br>(0.000, 0.308) |
| Erythema | 0.000<br>(0.000, 0.308) | 0.000<br>(0.000, 0.308) | 0.000<br>(0.000, 0.308) |
| Induration | 0.000<br>(0.000, 0.308) | 0.000<br>(0.000, 0.308) | 0.000<br>(0.000, 0.308) |
| Edema | 0.000<br>(0.000, 0.308) | 0.000<br>(0.000, 0.308) | 0.000<br>(0.000, 0.308) |
| Local pain | 0.900<br>(0.555, 0.997) | 0.556<br>(0.212, 0.863) | 0.400<br>(0.122, 0.738) |
| Involuntary muscle contractions,<br>light | 0.100<br>(0.003, 0.445) | 0.000<br>(0.000, 0.308) | 0.000<br>(0.000, 0.308) |

\*One missing value for local pain

With regards to systemic adverse events, two participants had fever at D7, and two others had light local hyperemia and headache at D37, probably related to the vaccine. Physical exams on all days (baseline, 0, 7, 30, 37, 60, 67, 90, and 120) showed no clinical alterations or abnormalities. Laboratory exams did not show alterations that could be classified as toxicity according to the ICTDR criteria, except for ALT and GGT, with few participants showing elevated values within toxicity levels. However, these participants also had elevated values prior to the vaccination (at baseline), and since they had no other abnormal laboratory findings or clinical symptoms, they were considered as most probably not related to the vaccine. All other single instances of values outside of the normal range were evaluated and, given that no abnormal laboratory findings or clinical symptoms were observed, they were all considered as non-clinically significant (that is, most probably not related to the vaccine). These data show that the vaccine produced very few adverse effects and all the observed events were local and mild (Tables 3 and 4).

**Table 3.** Secondary adverse events* in Sm14 vaccinated young.

| Day of Exam | Symptoms | Relationship to the Sm14 Vaccine |
| --- | --- | --- |
| D0** | Patient #10: Vomit, light.<br>Patient #13: Sleepiness, light. | Not related to the vaccine. |
| D7 | Patient #7: Fever.<br>Patient #8: Fever. | Fever in patients #7 and #8:<br>probably related to vaccine |
| D30 | Patient #12: Gallstones, light | Not related to the vaccine. |
| D37 | Patient #10: Local hyperemia light.<br>Patient #13: Light hepatic steatosis;<br>light headache. | Patient #10 probably related to<br>vaccine<br>#13 possibly related to vaccine. |
| D60 | Patient #3: Cold.<br>Patient #10: Flu, light.<br>Patient #14: Urinary tract infection,<br>light | Not related to the vaccine |
| D67 | Patient #1: Cervicalgia, light.<br>Patient #5: Pain in right heel, light. | Not related to the vaccine |
| D90 | Patient #7: Vaginal discharge, light.<br>Patient #13: Flu, light. | Not related to the vaccine |
| D120 | Patient #2: Light elevation of GGT.<br>Patient #3: Moderate eosinophilia.<br>Patient #10: Light Headache. | Not related to the vaccine |

\* New symptoms since last examen.
\*\* Previous exam was at Baseline. D0 is the day of the first dose.
Numbers preceded by # correspond to participant's code numbers.

**Table 4.**
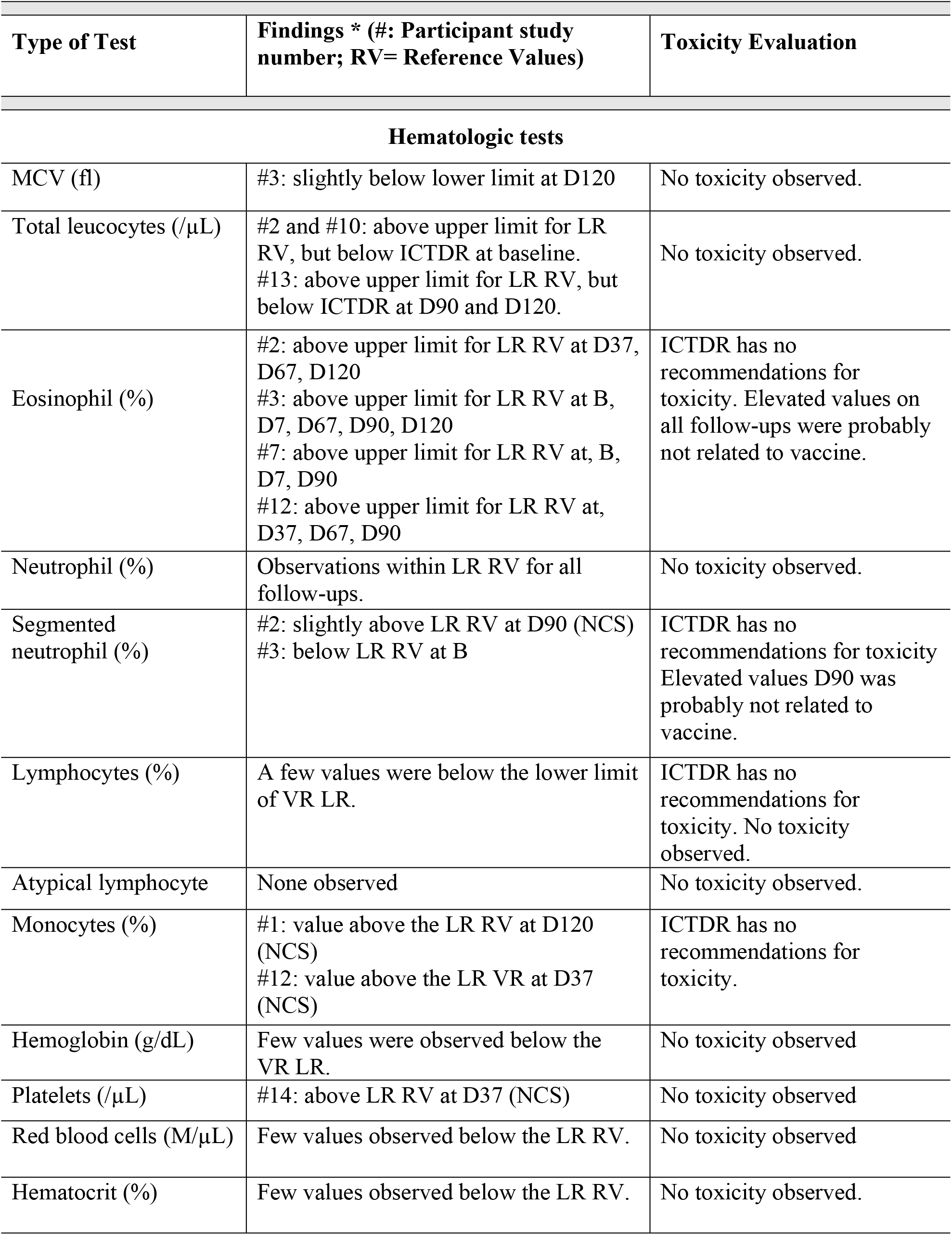
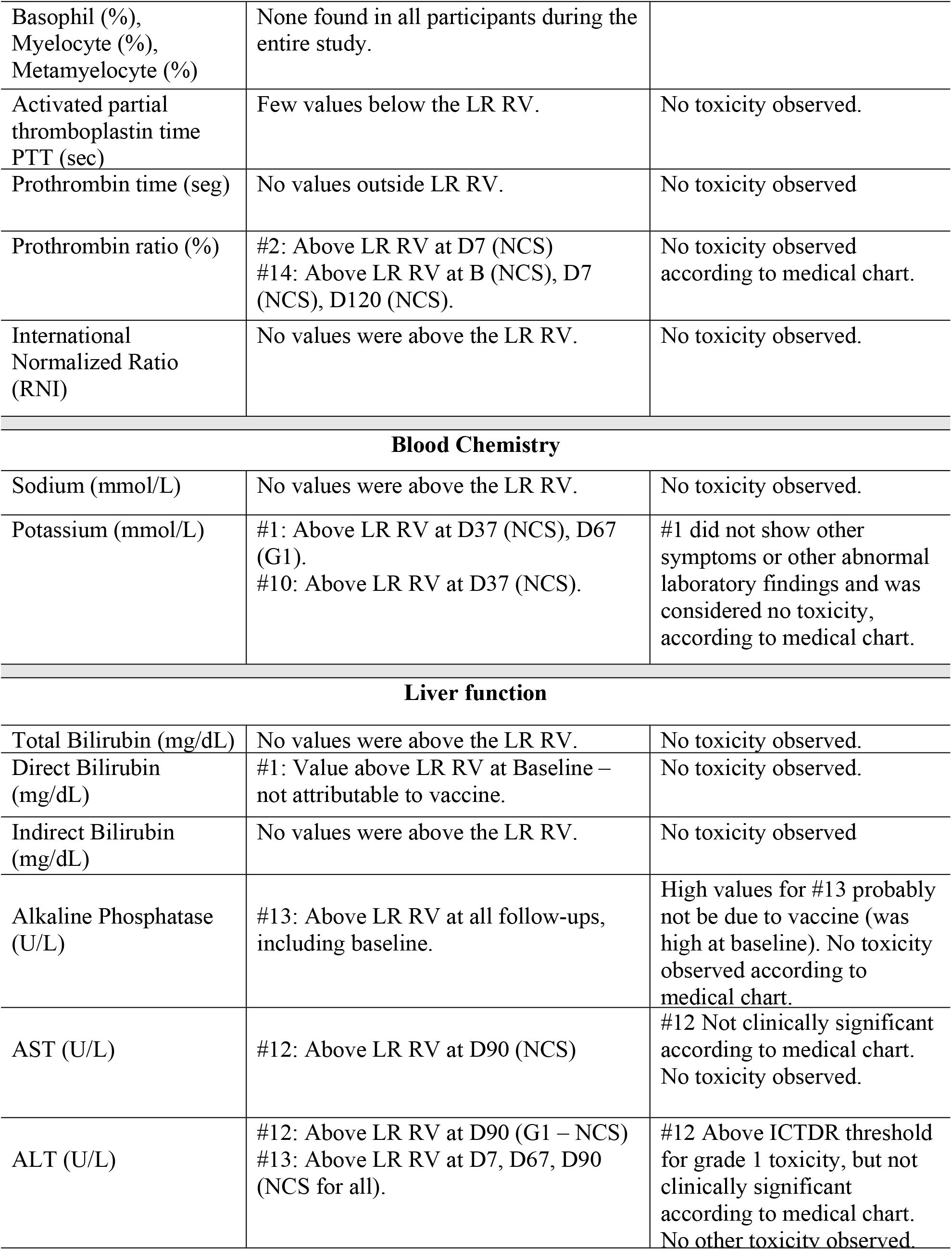
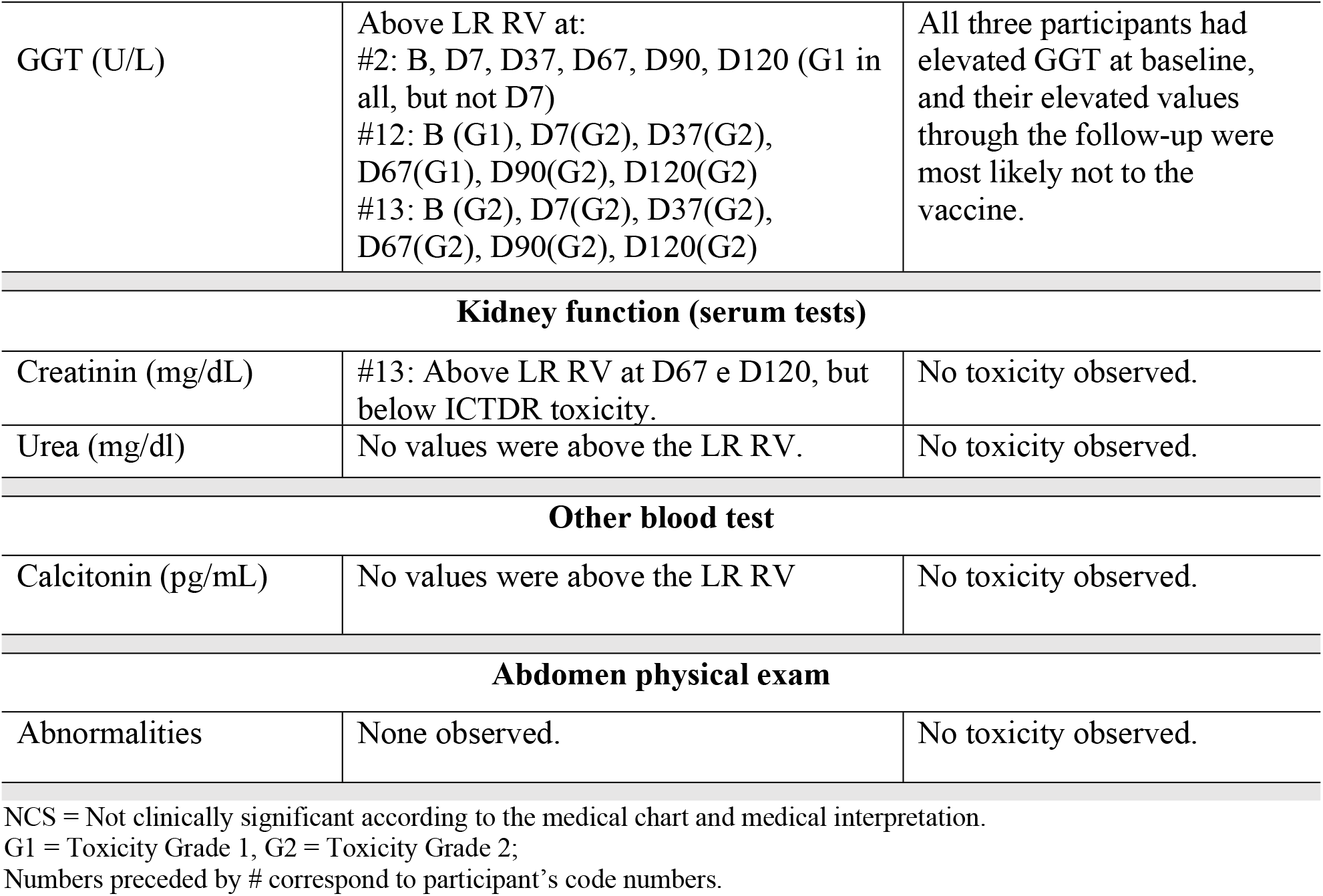
Summary of laboratory tests and physical exams in Sm14 vaccinated young adult women.

| Type of Test | Findings * (#: Participant study number; RV= Reference Values) | Toxicity Evaluation |
| --- | --- | --- |
| **Hematologic tests** |  |  |
| MCV (fl) | #3: slightly below lower limit at D120 | No toxicity observed. |
| Total leucocytes (/μL) | #2 and #10: above upper limit for LR RV, but below ICTDR at baseline.<br>#13: above upper limit for LR RV, but below ICTDR at D90 and D120. | No toxicity observed. |
| Eosinophil (%) | #2: above upper limit for LR RV at D37, D67, D120<br>#3: above upper limit for LR RV at B, D7, D67, D90, D120<br>#7: above upper limit for LR RV at, B, D7, D90<br>#12: above upper limit for LR RV at, D37, D67, D90 | ICTDR has no recommendations for toxicity. Elevated values on all follow-ups were probably not related to vaccine. |
| Neutrophil (%) | Observations within LR RV for all follow-ups. | No toxicity observed. |
| Segmented neutrophil (%) | #2: slightly above LR RV at D90 (NCS)<br>#3: below LR RV at B | ICTDR has no recommendations for toxicity<br>Elevated values D90 was probably not related to vaccine. |
| Lymphocytes (%) | A few values were below the lower limit of VR LR. | ICTDR has no recommendations for toxicity. No toxicity observed. |
| Atypical lymphocyte | None observed | No toxicity observed. |
| Monocytes (%) | #1: value above the LR RV at D120 (NCS)<br>#12: value above the LR VR at D37 (NCS) | ICTDR has no recommendations for toxicity. |
| Hemoglobin (g/dL) | Few values were observed below the VR LR. | No toxicity observed |
| Platelets (/μL) | #14: above LR RV at D37 (NCS) | No toxicity observed |
| Red blood cells (M/μL) | Few values observed below the LR RV. | No toxicity observed |
| Hematocrit (%) | Few values observed below the LR RV. | No toxicity observed. |
| Basophil (%),<br>Myelocyte (%),<br>Metamyelocyte (%) | None found in all participants during the entire study. |  |
| Activated partial thromboplastin time PTT (sec) | Few values below the LR RV. | No toxicity observed. |
| Prothrombin time (seg) | No values outside LR RV. | No toxicity observed |
| Prothrombin ratio (%) | #2: Above LR RV at D7 (NCS)<br>#14: Above LR RV at B (NCS), D7 (NCS), D120 (NCS). | No toxicity observed according to medical chart. |
| International Normalized Ratio (RNI) | No values were above the LR RV. | No toxicity observed. |
| Blood Chemistry |  |  |
| Sodium (mmol/L) | No values were above the LR RV. | No toxicity observed. |
| Potassium (mmol/L) | #1: Above LR RV at D37 (NCS), D67 (G1).<br>#10: Above LR RV at D37 (NCS). | #1 did not show other symptoms or other abnormal laboratory findings and was considered no toxicity, according to medical chart. |
| Liver function |  |  |
| Total Bilirubin (mg/dL) | No values were above the LR RV. | No toxicity observed. |
| Direct Bilirubin (mg/dL) | #1: Value above LR RV at Baseline – not attributable to vaccine. | No toxicity observed. |
| Indirect Bilirubin (mg/dL) | No values were above the LR RV. | No toxicity observed |
| Alkaline Phosphatase (U/L) | #13: Above LR RV at all follow-ups, including baseline. | High values for #13 probably not be due to vaccine (was high at baseline). No toxicity observed according to medical chart. |
| AST (U/L) | #12: Above LR RV at D90 (NCS) | #12 Not clinically significant according to medical chart. No toxicity observed. |
| ALT (U/L) | #12: Above LR RV at D90 (G1 – NCS)<br>#13: Above LR RV at D7, D67, D90 (NCS for all). | #12 Above ICTDR threshold for grade 1 toxicity, but not clinically significant according to medical chart. No other toxicity observed. |
| GGT (U/L) | Above LR RV at:<br>#2: B, D7, D37, D67, D90, D120 (G1 in all, but not D7)<br>#12: B (G1), D7(G2), D37(G2), D67(G1), D90(G2), D120(G2)<br>#13: B (G2), D7(G2), D37(G2), D67(G2), D90(G2), D120(G2) | All three participants had elevated GGT at baseline, and their elevated values through the follow-up were most likely not to the vaccine. |
| **Kidney function (serum tests)** |  |  |
| Creatinin (mg/dL) | #13: Above LR RV at D67 e D120, but below ICTDR toxicity. | No toxicity observed. |
| Urea (mg/dl) | No values were above the LR RV. | No toxicity observed. |
| **Other blood test** |  |  |
| Calcitonin (pg/mL) | No values were above the LR RV | No toxicity observed. |
| **Abdomen physical exam** |  |  |
| Abnormalities | None observed. | No toxicity observed. |

NCS = Not clinically significant according to the medical chart and medical interpretation.
G1 = Toxicity Grade 1, G2 = Toxicity Grade 2;
Numbers preceded by # correspond to participant's code numbers.

### Sm14 does induce adaptive immune responses in healthy women

As occurred with male subjects on Phase IA [22], the immune response of females vaccinated with the Sm14+GLA-SE was characterized by high levels of anti-Sm14 IgG, mainly of the IgG_1_ isotype. Responses were augmented after the second dose. Ninety percent of vaccinated subjects developed higher specific total IgG antibody titres 90 days post vaccination and theses titres remained as high up to 120 days. Compared to baseline, total specific IgG, as well as IgG_1_ and IgG_3_, increases were statistically significant from day-60 (Figure 3). The levels of specific IgG_2_ and IgG_4_ increased after the second dose and achieved statistical significance from day-90 up to day-120. Also, neither general nor specific IgE increases were detected at any time.

**Figure 3.**
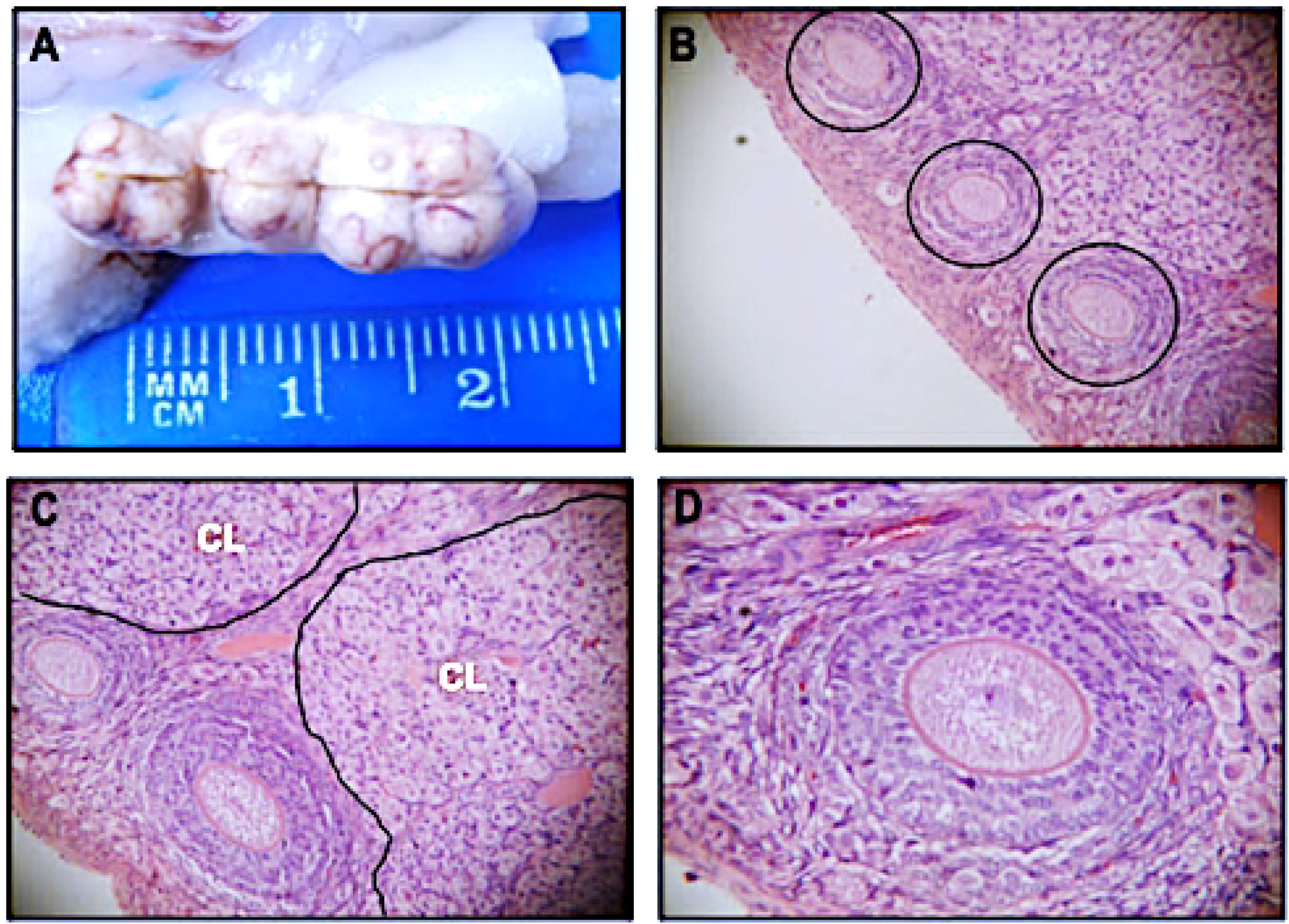
Levels of anti-Sm14 antibody classes and cytokine production stimulated by vaccination. (A) ELISA antibody results (B) Percentage of CD4^+^ T cells expressing combinations of IFNγ, TNF_α_ and IL-2 production in CD4+ T cells on days 0, 30, 90 and 120. *p*-values for comparison was performed for vaccinated group to baseline (Day 0). \**p*-values at the 0.05 significance level.

The Sm14+GLA-SE vaccination elicited robust cytokine responses. Cytokine profiles show increased TNFα, IFNγ, and IL-2 in vaccinated individuals on days 90 and 120. Analysis results indicated overall comparable responses among male and female cohorts [22] (Fig 3).

## Discussion

Safety is the most important attribute of a vaccine designed for large-scale use in endemic populations. The studies described herein were the basis for legal authorization to start clinical trials for the Sm14 vaccine in Brazil. The preparation was evaluated according to WHO recommendations and guidelines in multiple-dose and reproductive studies that provided pre-clinical data ensuring the safety of the vaccine formulation [34].

For obtaining authorization from the Brazilian health authorities to initiate phase I clinical trials of Sm14+GLA-SE anti-schistosomiasis vaccine, Anvisa has approved the study to be performed in healthy men living in non-endemic areas for schistosomiasis but did impose a restriction to the engagement of women of childbearing age. A preambular reproductive toxicity test was required in pregnant rabbits, to ensure vaccine safety for this population group. The variations observed in laboratory blood tests did not indicate a significant effect on the health of the animals. Coherently, the animals did not show relevant clinical alterations associated with the study and no macroscopic or histopathological alterations were observed that represented clinical significance. Most of the analysed parameters were within the normal range of variation, indicating that the vaccine was safe and well tolerated at either dose level, in addition to providing clear humoral immune response.

These data provided the green light to the Sm14 vaccine clinical test in women at childbearing age, to evaluate the clinical steps of the safety and immunologic evaluation of the Sm14+GLA-SE anti-schistosomiasis candidate vaccine in healthy volunteer women (clinical trial Phase Ib).

Results obtained in women have confirmed the pattern of responses of Phase Ia with male adults [22]. Our immunological analysis revealed that Sm14 induced a broad spectrum of immune responses stimulating both Th1 and Th2 cytokines However, there was neither a specific nor a general IgE increase, an immunoglobulin associated to the pathogenesis of immediate hypersensitivity reactions because of its ability to bind specifically to high affinity receptors on mast cells or basophils by the alpha chain of the FC receptor.

## Conclusion and perspectives

Preclinical assays on pregnant rabbits revealed that the candidate anti-schistosomiasis rSm14-GLA-SE vaccine test resulted in no changes concerning the reproductive status and well as other pregnancy parameters. Moreover, Phase Ib clinical trial applied to healthy young adult women confirmed that no serious adverse events were observed after three doses of 50 µg Sm14 + 10µg GLA-SE in 10 women from a schistosomiasis non-endemic area, administered three times with 30-day interval.

Based on data generated from the clinical studies done so far, severe toxicity (Grade 3 or 4), either local and/or systemic, is not expected to occur. The administration of the investigational product is anticipated to be well tolerated. So, local reactions (such as pain at the site of injection, erythema, induration, edema, or pruritus) may occur but are usually mild, self-limited, and subside without treatment.

As observed on this trial, adaptive cell-mediated and humoral immune responses (IgG and isotypes, but not IgE), along with many previous preclinical results, strongly support Sm14 as an effective vaccine against *S. mansoni* infection. Possibly, it will also be useful against infections caused by other species of *Schistosoma*.

Clinical studies are a prerequisite for licensing drugs and immunobiologicals by regulatory agencies in most countries. In this way, they are a constituent part of the process of research and development of supplies for human health. The Sm14+GLA-SE vaccine is at an advanced stage of development, after successfully passing through pre-clinical stages and Clinical phases Ia and Ib, is presently in phase II, clinical trial in the Senegal River Delta region, in African endemic sites for both *S. mansoni* and *S. haemathobium*. It is believed that the development of a safe, effective, and humanitarian vaccine will change the epidemiological profile of this disease, thus freeing countries such as Brazil and much of the African continent from one of the major stigmas of underdevelopment, which are parasitic diseases and particularly schistosomiasis.

## Data Availability

All related data are included in the manuscript or in the supplementary files.

## Acknowledgements

We thank the researchers and nurses from the staff of the INI/Fiocruz, for conducting the study. Lots of thanks to Tom Vedvick of the Infectious Disease Research Institute, for most valuable advices and support for the formulation of Sm14with GLA, to Andrew J. G. Simpson, scientific director of the company Orygen Biotecnologia S.A, that funded part of clinical trials for the Sm14-GLA-SE Schistosomiasis Vaccine Development Project, to Donald De Roo, from LICR for technical support for setting up the Fill Finish procedures at Florida Biologix and Quality Control Panel at PPD Inc., to Carlos Henrique Henrique of Ourofino Animal Health Inc. for support at all levels with operational steps and pre-clinical animal tests and finally to Jorge Bermudez, former Vice President of Innovation and Production of Fiocruz and to PHI/WHO(World Health Organization) for strategic support.

## Supporting information

**S1 Table. Demographic characteristics of the female participants at the baseline**.

(DOCX)

**S1 Fig. Flowchart for the phase 1b clinical trial, including enrollment and clinical follow-up at the Brazilian National Institute of Infectology Evandro Chagas**.

(TIF)

